# Predicting 30-Day Hospital Readmission in Medicare Patients: Insights from an LSTM Deep Learning Model

**DOI:** 10.1101/2024.09.08.24313212

**Authors:** Xintao Li, Sibei Liu

## Abstract

**Background:** Readmissions among Medicare beneficiaries are a major problem for the US healthcare system from a perspective of both healthcare operations and patient caregiving outcomes. Our study analyzes Medicare hospital readmissions using LSTM networks with feature engineering to assess feature contributions.

**Design:** The 21002 senior patient admission data from MIMIC-III clinical database at Beth Israel Deaconess Medical Center.is utilized in the study We selected variables from admission-level data, inpatient medical history and patient demography. The baseline model is a logistic-regression model based on the LACE index, and the LSTM model is designed to capture temporal dynamic in the data from admission-level and patient-level data. We leveraged Area Under the Curve metric, precision and recall to evaluate the model’s performance.

**Results:** The LSTM model outperformed the logistic regression baseline, accurately leveraging temporal features to predict readmission. The major features were the Charlson Comorbidity Index, hospital length of stay, the hospital admissions over the past 6 months or the number of medications before discharge, while demographic variables were less impactful

**Limitations:** The use of a single-center database from the MIMIC-III database limits the generalizability of the findings. Additionally, the exclusion for specific chronic conditions and external factors limit the model’s ability to capture the complexities of chronic diseases.

**Conclusions:** This work suggests that LSTM networks offers a more promising approach to improve Medicare patient readmission prediction. It captures temporal interactions in patient databases, enhancing current prediction models for healthcare providers.

**Implications:** Adoption of predictive models into clinical practice may be more effective in identifying Medicare patients to provide early and targeted interventions to improve patient outcomes.

**Highlights:**

- **Improved Prediction:** Our LSTM model outperforms the logistic regression model with LACE index in predicting Medicare patient readmissions.
- **Feature Contribution:** Feature engineering ranks variables base on the impact, deprioritizing the importance of patient demographic variables, highlighting the importance of patients’ chronic diseases in leading hospitalizations and guiding targeted interventions to prevent senior hospital readmissions for healthcare providers.
- **Effective Use of Data:** Our LSTM model incorporates with time-series data from MIMIC-III database to enhance the accuracy of all-cause hospital readmission predictions, especially for the high-risk patients.
- **Actionable Insights:** The result demonstrates the utilization of deep learning model in healthcare decision-making to reduce hospital readmissions for seniors.

## Introduction

Medicare beneficiary readmissions is one of the major challenges in US healthcare system from a perspective of both healthcare operations and patient caregiving outcomes^1^. The Hospital Readmissions Reduction Program (HRRP)^2^ created by the US federal Centers for Medicare & Medicaid Services (CMS) serves as a major initiative of penalizing excess 30-day readmissions to the hospitals. The prediction and prevention of senior readmission are now becoming major goals of improving patient outcomes and reducing spending on medical care. The features related to senior readmission provides valuable information of the insight to healthcare provider to optimize the medical decision on the prevention.

There have been various studies focusing on the prediction of hospital readmission, which target specific subpopulations to enhance predictive accuracy. For example, a study on diabetic patients^3^ utilized a deep learning model combining wavelet transform and deep forest techniques, achieving an AUROC of 0.726. Similarly, research involving heart failure patients employed random forests with administrative claims data, resulting in an AUC above 0.800. Having said this, even though studies with potentially higher predictive accuracy have been published, the LACE index^4^ remains the best-known model for predicting readmissions for general patient populations. Validated through a large-scale study of 1,000,000 Ontarians discharged from hospitals, the LACE index incorporates variables such as length of stay, acuity of admission, comorbidity, and emergency department use, demonstrating solid predictive performance with a C statistic of 0.684.

Significant limitations persist regarding the variables typically used in readmission prediction. While medical comorbidities and pre-existing predictive scores, such as the Charlson Comorbidity Index^5^, are frequently included, basic sociodemographic variables like age and gender do not consistently enhance model performance. Also often ignored are variables related to interventions during the admission and the post-discharge period^6^, which are important for prediction but hard to include due to data constraints. Another critical gap in current research is the interpretability of predictive models. Traditional models frequently fail to leverage the sequential and time-series data inherent in electronic health records (EHRs)^7,8^, potentially leading to information loss. Moreover, the deep learning models take decision-making to a black-box level, which is challenging to use in a clinical environment where model reliability and interpretability are important^9–14^.

This study aims to develop a comprehensive deep learning model to address the challenge of predicting hospital readmissions within 30 days after discharge, specifically targeting senior Medicare patients. Our approach introduces innovative strategies in data extraction and variable selection, coupled with a recurrent neural network (RNN) architecture with long short-term memory (LSTM) layers that effectively leverages time-series data. To prevent overfitting, we deprioritize patient demographic variables, as they offer minimal contribution to model performance. Instead, our model integrates admission-level data, such as surgeries, consultations, and medications, with patient-level data before and after discharge, including ICD codes and ED admission history. The LSTM model’s performance is compared with a logistic regression model incorporating the LACE index risk score, allowing us to compare its effectiveness in predicting readmissions among Medicare patients.

## Methods

### Data Collection

The dataset for this study was sourced from the MIMIC-III database^15^, which is a comprehensive, single-center resource containing detailed clinical information of patients admitted to critical care units at Beth Israel Deaconess Medical Center in Boston, Massachusetts. MIMIC-III comprises data for 53,423 unique hospital admissions of adult patients (aged 16 years and above) between 2001 and 2012. This data source has been widely used to train various machine learning models, such as Random Forest, XGBoost^16,17^, and contributed to a huge improvement in the healthcare research area.

The data processing flow is showed in **Figure 1**. Initially, we ensured the integrity of the dataset by addressing anomalies such as implausible ages (e.g., >120 years) and missing values. Our focus was narrowed to 21,002 Medicare patient admissions from a total of 46,520, utilizing their demographic details. Furthermore, we analyzed 6,984 distinct ICD codes to account for the patients’ medical histories. This robust dataset serves as the foundation for developing and validating our predictive model.

**Figure 1.**
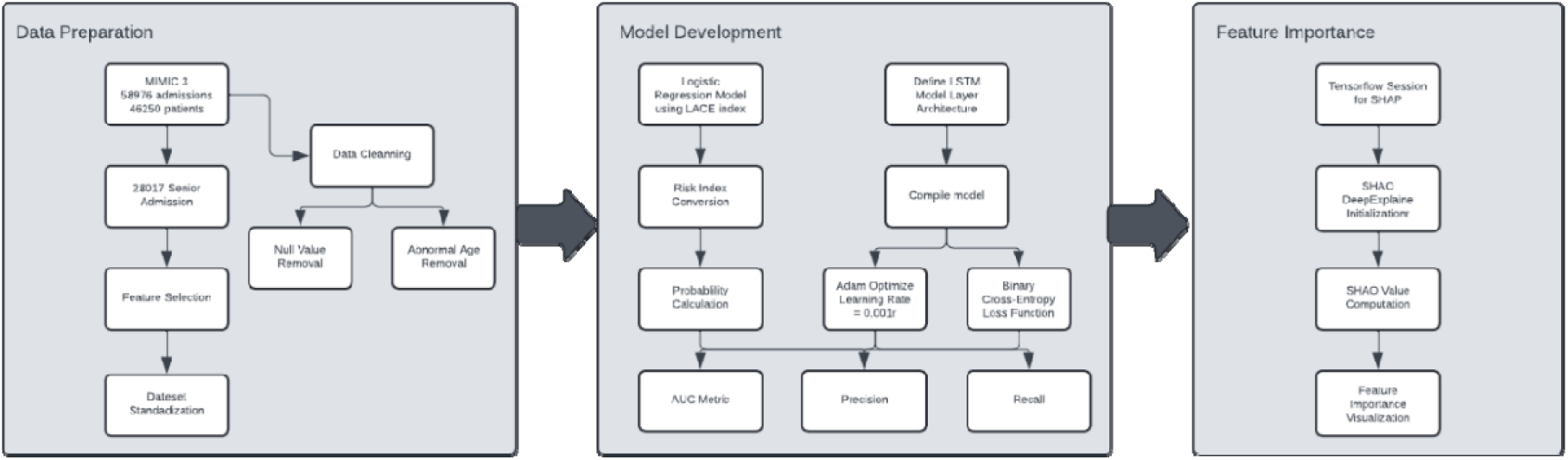
Flow Chart of Data Preparation, Model Development and Feature Importance Analysis

### Feature Extraction

We selected variables based on significant tests and evaluations of in-hospital treatments for elderly patients, as discussed in Taylor’s research^18^. The extracted features are grouped into three categories: admission-level data, medical history data, and patient demographic data.

### Admission-Level Data

This data is crucial for modeling hospital readmission risk, capturing the interventions and medical treatments received during the hospital stay^19,20^. Key features include:

- **Emergent Admission:** Identifies whether the hospital admission was acute or unplanned.
- **Surgery During Admission:** The count of surgical procedures performed during the hospital stay.
- **Length of Stay (LOS):** The duration of the hospital stay, with longer stays correlating with an increased likelihood of readmission^21^, as adjusted for other.
- **Medications:** The number of medications prescribed before discharge.
- **Consultations:** Indicates whether thorough medical evaluations, including Echo, ECG, and radiology reports, were conducted before discharge.
- **Seasons:** Encodes seasonal variations, as temperature fluctuations significantly impact hospital admission rates^22^.

### Medical History Data

Research indicates that 29% of hospitalized patients are readmitted post-discharge in all-cause admission studies^23^. For this study, we extracted:

- **Past 6 Months Hospital Admission:** The number of hospital admissions within the past six months.
- **Past 6 Months ED Admission:** The number of emergency department admissions within the past six months.
- **Charlson Comorbidity Index Score:** A weighted score evaluating the severity of comorbid diseases, a critical factor associated with readmissions^5^.

### Demographic Data

Given the reduced emphasis on demographic factors, only age and gender—recognized as the most significant demographic variables—were included in the model’s training. This comprehensive feature extraction strategy ensures that our model accounts for crucial factors influencing hospital readmissions, particularly for elderly patients, while mitigating overfitting by deprioritizing less impactful variables^24,25^. All variables we used in our model are listed in **Table 1**.

**Table 1.** Patients Characteristics and 30-Day Hospital Readmission.

| Characteristic | Category/Value | Number (%) of patients<br>n = 20186 | Readmission Within 30 Days<br>After Discharge<br>Number (%) Of Patients |  |
| --- | --- | --- | --- | --- |
|  |  |  | No<br>n = 18891<br>(93.8) | Yes<br>n = 1295 (6.2) |
| **Admission-Level Data** |  |  |  |  |
| **Emergent Admission** | Yes | 16986 (84.1) | 15858 (83.9) | 1128 (87.1) |
|  | No | 3200 (15.9) | 3033 (16.1) | 167 (12.9) |
| **Surgery During Admission** | Yes | 454 (2.2) | 419 (2.2) | 35 (2.7) |
|  | No | 19732 (97.8) | 18472 (97.8) | 1260 (97.3) |
| **Length Of Stay (Los)** | Median (IQR) | 7 (4-12) | 7 (4-12) | 8 (5-14) |
| **Medications** | Median (IQR) | 39 (26-55) | 39 (25-55) | 40 (28-56) |
| **Consultations** | Median (IQR) | 17 (9-32) | 16 (9-31) | 19 (10-41) |
| **Seasons** | Spring | 4908 (24.3) | 4606 (24.4) | 302 (23.3) |
|  | Summer | 5198 (25.8) | 4882 (25.8) | 316 (24.4) |
|  | Autumn | 5145 (25.5) | 4788 (25.3) | 357 (27.6) |
|  | Winter | 4935 (24.4) | 4615 (24.4) | 320 (24.7) |
| **Medical History Data** |  |  |  |  |
| **Past 6 Months Hospital Admission** | Median (IQR) | 0 (0-0) | 0 (0-0) | 0 (0-1) |
|  | >0 | 2605 (12.9) | 2273 (12.0) | 332 (25.6) |
| **Past 6 Months Ed Admission** | Median (IQR) | 1 (0-1) | 1 (0-1) | 1 (0-1) |
|  | >0 | 11932 (59.1) | 11055 (58.5) | 877 (67.7) |
| **Charlson Comorbidity Index Score** | Median (IQR) | 3 (1-6) | 3 (1-6) | 6 (3-10) |
| **Demographic Data** |  |  |  |  |
| **Gender** | Female | 9495 (47.0) | 8935 (47.3) | 560 (43.2) |
|  | Male | 10691 (53.0) | 9956 (52.7) | 735 (56.8) |
| **Age** | Median (IQR) | 77 (71-82) | 77 (71-82) | 77 (70-82) |

### Model Development

#### Baseline Model: LACE Index

To get a robust baseline model for senior readmission prediction, we developed a multidimension logistic regression model with LACE index components^4^. Logistic regression is the statistical methods for that models the binary outcomes of one or more independent variables. The logistic regression model is of the form as:

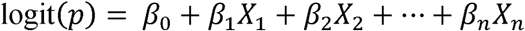

In this equation, *p* represents the probability of readmission, *β*_0_ is the intercept, and *β*_i_are the coefficients corresponding to the predictor variables *X_i_*. The LACE index components—Length of stay (L), Acuity of the admission (A), Comorbidities (C), and Emergency department visits (E)—serve as the independent variables in our model. The definition of LACE index is listed in Table 2.

**Table 2.** Components and Scoring of the LACE Index for 30-Day Readmission Risk.

| **LACE Component** | **Criteria** | **Score** |
| --- | --- | --- |
| **Length Of Stay (L)** | < 1 | 0 |
|  | 1 | 1 |
|  | 2 | 2 |
|  | 3 | 3 |
|  | 4-6 | 4 |
|  | 7-13 | 6 |
|  | >14 | 7 |
| **Acuity of Admission (A)** | Yes | 3 |
| **Charlson Comorbidity Index (C)** | 0 | 0 |
|  | 1 | 1 |
|  | 2 | 2 |
|  | 3 | 3 |
|  | >=4 | 5 |
| **Visits To Emergency Department (E) In Previous 6 Months** | 0 | 0 |
|  | 1 | 1 |
|  | 2 | 2 |
|  | 3 | 3 |
| | $\geq 4$ | 4 |

To enhance the interpretability of the model, we adopted the approach suggested by Sullivan et al.^26^ to transform the logistic regression model into a risk index. This step involves converting a model’s regression coefficients into point scores by taking each coefficient and dividing by the smallest absolute value of any coefficient in the model, and then rounding the result off to the nearest whole number. The overall risk score for an individual patient is the total of those points. The LACE index components consist of Length of stay (L), Acuity of the admission (A), Comorbidities (C), and Emergency department visits (E), which serve as the independent variables in our model.

The probability of readmission is subsequently estimated using the formula:

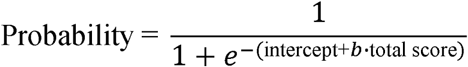

where *b* is the coefficient in the regression model with the smallest absolute value.

The logistic regression model was trained and validated using a 70-30 train-test split, allowing us to provide a benchmark for more complex (and hopefully better) models that could be trained by using deep learning techniques.

#### Long short-term memory (LSTM) model

We utilized Long Short-Term Memory (LSTM) networks to build our predictive model on time series data^27^. The clinical data from patients are usually with irregular intervals and missing-value imputation (end-diastole, end-systole, COK time intervals) that requires special handling^28^. The LSTM networks were particularly a good fit for temporal dependencies and patterns^29^. LSTM networks is a specialized form of Recurrent Neural Networks (RNNs) that designed to manage long-term dependencies in sequential data(). The unique architecture composed of a series of gates: forget, input, and output gates, which regulate the information flow and ensure that relevant details are retained over extended periods^28^. The LSTM is of the form as

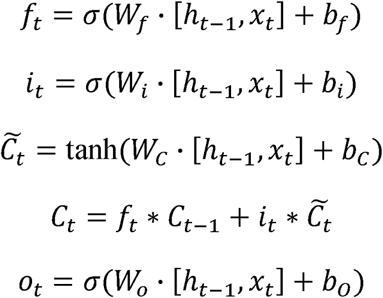

Here, *f_t_* is the forget gate, *i_t_* is the input gate, *c̃_t_* is the candidate cell state, *C_t_* is the cell state, *o_t_* is the output gate, and *h_t_* is the hidden state. The sigmoid function (σ) and hyperbolic tangent function (tanh) introduce non-linearity to the model^30^.

Our LSTM model architecture consists of a bidirectional LSTM layer and an additional LSTM layer, allowing for the model to adapt to patterns in both forwards and backwards directions in the time-series data^28^. Because temporal dynamics can be complex in many situations, dual-layer LSTMs are better at accommodating nonlinear interactions between how future and past events influence each other, especially when we want to rely on how well our model is able to predict several future steps. The decimal forecast was assigned to the single output neuron in a dense decision layer, activated by a sigmoid function, which is used for binary classification^31^. The sigmoid function that maps input values between 0 and 1 as:

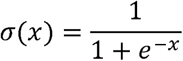

Figure 2 and Figure 3 illustrate how LSTM does work on top of RNN. Figure 2 is a sequential model of LSTM. The sequential model can work on each time step until it reaches the end of the data. Every step of the process has input that can influence the short term and long-term memory, while forget, input, and output gates to regulate the information flow^32,33^. The interplay between these gates ensures that the LSTM can selectively remember and utilize relevant information over long sequences. This is crucial for tasks of time-series data or sequential information processing of any form^34,35^. Figure 3 exhibits how in each time step, it depends on the others, so this helps the network to capture some complicated temporal pattern in the sequence data. By remembering and update the long-term memory state of the dependency through time flow, the LSTM can avoid the issue of vanishing gradient, which occurs in a traditional RNN^36^. Thus, LSTM provides a robust way to manage the long-term dependency in the input data.

**Figure 2.**
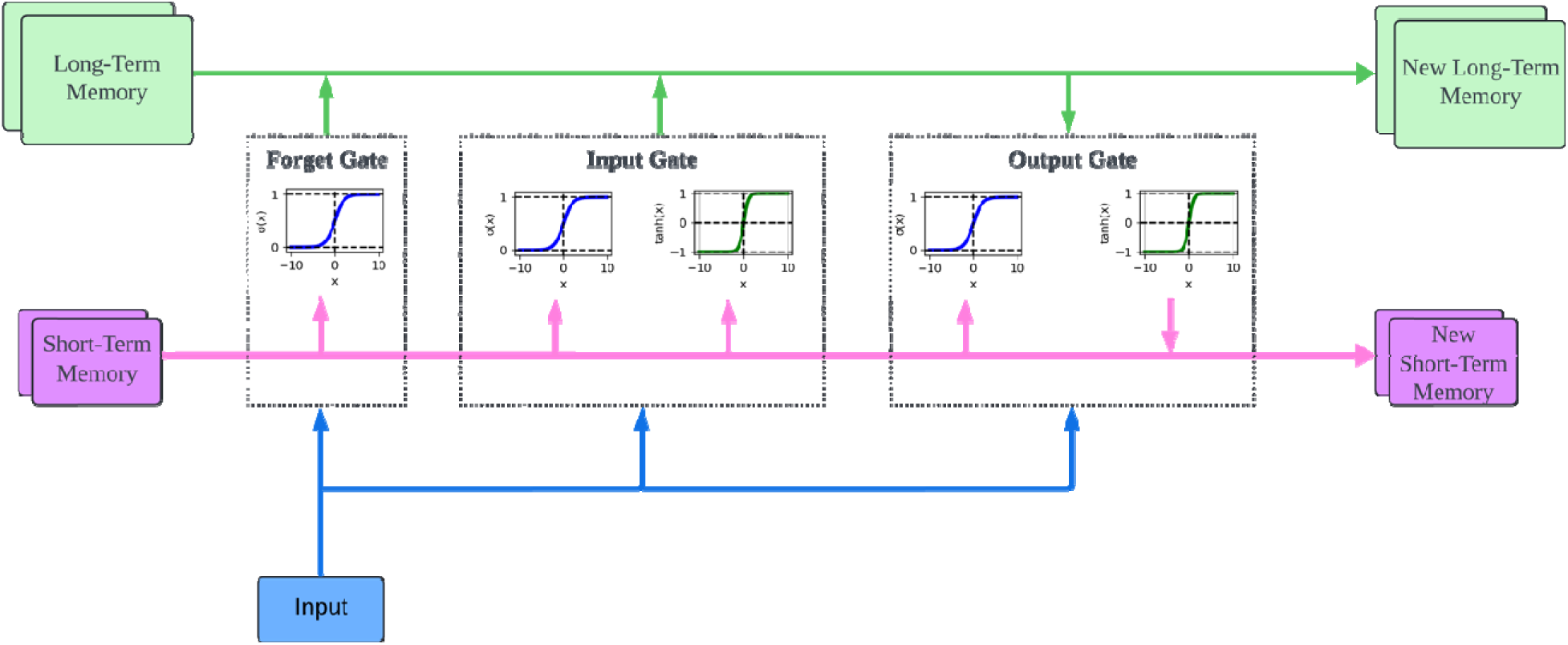
Long Short-Term Memory Neural Networks

**Figure 3.**
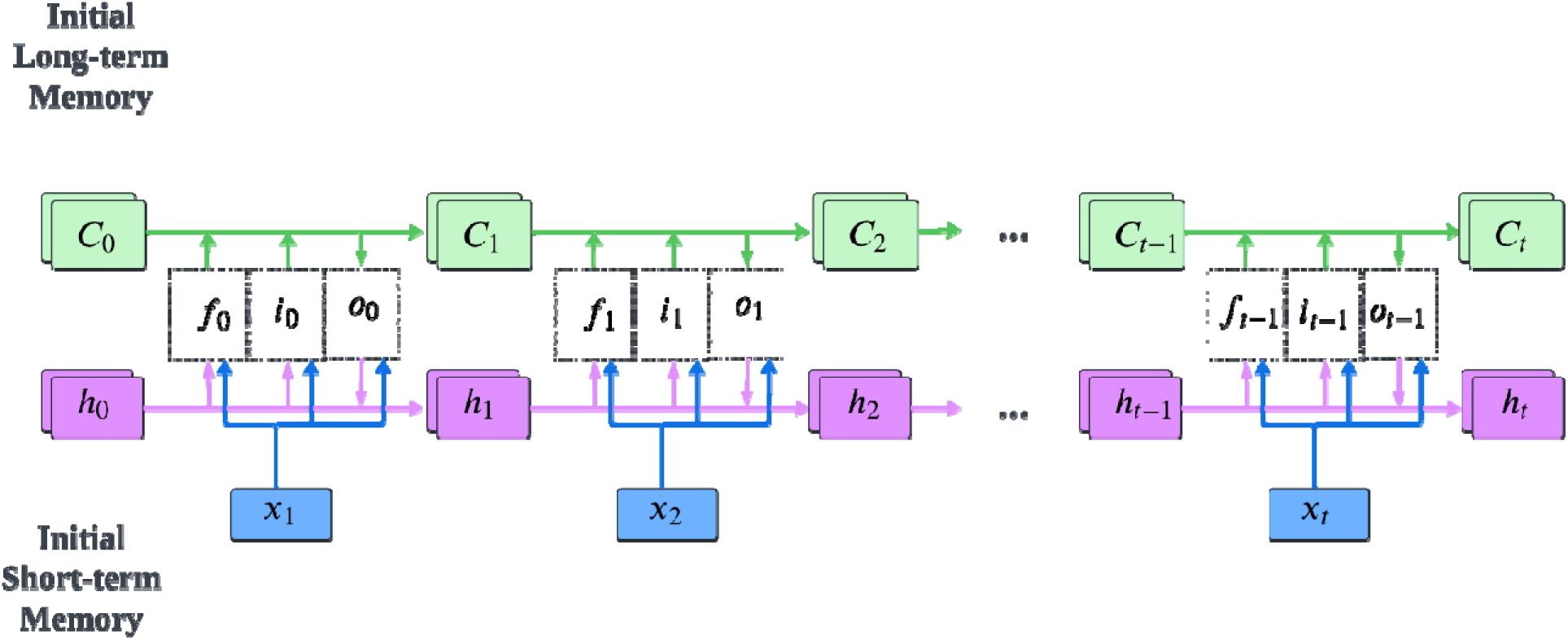
Long Short-Term Memory Flow Chart

**Figure 4.**
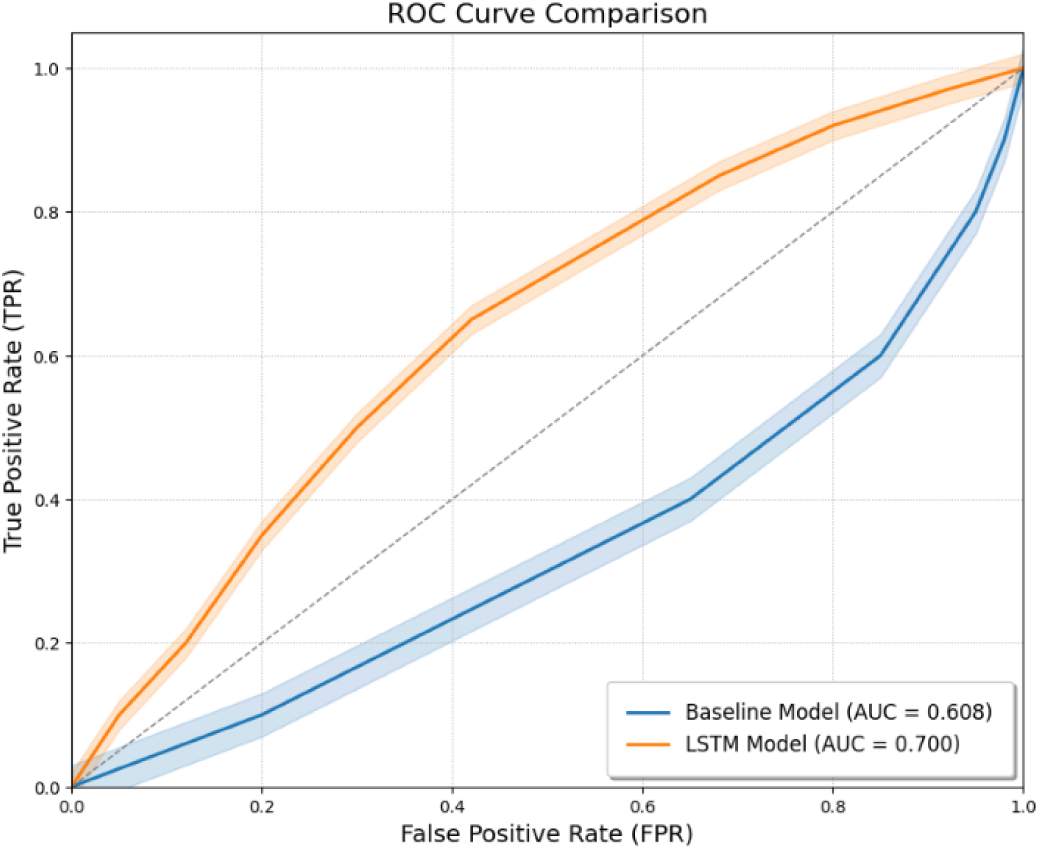
AUC Comparation of Logistic Regression and LSTM

We trained our model with 64 epochs to achieve the highest AUC. To optimize the model, we used the Adam optimizer with a learning rate of 0.001, selected through hyperparameter tuning^37^. The Adam optimizer works well on datasets with sparse gradients and noisy data like our clinical dataset. We used binary cross-entropy as the loss function to measure the discrepancy between the predicted and actual values:

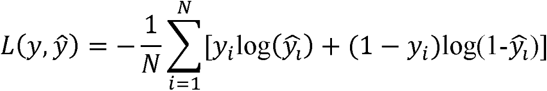

where y is the true label, ŷ is the predicted probability, and N is the number of samples. Additionally, we used the Area Under the Curve (AUC) metric to evaluate the model’s performance.

We utilized layer wise dropout on the deep learning model to avoid the common overfitting problem in deep learning models. The dropout rate is 0.4.^38^. Finally, since the challenge does not require a fixed number of epochs for training, we utilized the Early Stopping method, it keeps track of the validation loss over training iterations, and halts training if the validation loss does not improve after following a fixed number of epochs.

The LSTM model was trained using a 70-30 train-test split for robust evaluation. By comparing the performance of the LSTM model with the logistic regression baseline, we aimed to show the advantages of incorporating temporal dynamics into predictive modeling for healthcare application.

### Evaluation

To evaluate the performance of our models, we utilized several key metrics. The Area Under the Receiver Operating Characteristic Curve (AUC-ROC) serves as the primary measure of predictive accuracy. The AUC-ROC is a widely recognized metric that evaluates a model’s ability to distinguish between positive (readmitted) and negative (not readmitted) classes. A higher AUC indicates superior model performance, suggesting that the model is better at correctly classifying patients.

In addition to AUC-ROC, we focused on precision and recall within the "highest risk decile" of patients. This involved ranking patients according to their predicted risk scores and analyzing the top 10%—those deemed most at risk^39–41^. Precision in this context reflects the proportion of patients identified as high-risk who were actually readmitted, providing a measure of the model’s accuracy in predicting true positives. Recall, conversely, measures the proportion of actual readmissions captured within this high-risk group, indicating the model’s effectiveness in identifying the majority of true readmissions.

Precision is defined as:

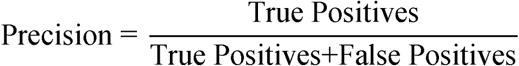

Recall is defined as:

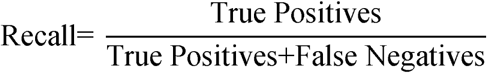

These metrics are critical for assessing the clinical utility of our models. High precision ensures that most patients flagged as high-risk are indeed readmitted, reducing unnecessary interventions. High recall, on the other hand, ensures that the model successfully identifies a substantial proportion of all actual readmissions, thereby minimizing missed cases^41^.

To ensure a robust and comprehensive evaluation, these metrics were averaged over 20 different data splits^42^. For each split, the data was divided into training (60%), validation (15%), and testing (25%) sets, with random assignments to reduce the potential for overfitting. This methodology provides a thorough comparison of model performance across different samples, ensuring that the evaluation is both reliable and generalizable.

### Results Baseline Model

The LACE index model, employing multivariable logistic regression, achieved an AUC of 0.608 (95% CI: 0.602 - 0.614). This performance is notably lower than the AUC of 0.85 reported when the LACE index is applied to a general population^43^. For high-risk patient readmission prediction, the model attained a precision of 0.168 (95% CI: 0.161 - 0.175) and a recall of 0.261 (95% CI: 0.252 - 0.270). This indicates that, on average, 16.8% of the patients predicted to be in the highest risk decile were indeed readmitted, while the model captured only 26.1% of the actual readmissions within this high-risk group.

Among the variables selected for this logistic regression model, several factors emerged as significant predictors of hospital readmission. Length of stay (OR 1.015, 95% CI: 1.009 - 1.018) was found to be a significant predictor, indicating that for each additional day spent in the hospital, the odds of readmission increase by approximately 1.5%. The Charlson comorbidity score (OR 1.127, 95% CI: 1.119 - 1.135) also significantly increased the likelihood of readmission, suggesting that patients with higher comorbidity burdens are at a greater risk. Additionally, past admissions within six months (OR 1.199, 95% CI: 1.054 - 1.324) were associated with a higher probability of readmission, underscoring the impact of recent healthcare utilization. Conversely, being female (PL_Sex) was associated with a reduced risk of readmission (OR 0.802, 95% CI: 0.731 - 0.867), indicating that females are at a higher risk compared to females. These findings highlight the importance of targeted interventions focusing on these significant predictors to reduce hospital readmissions.

### LSTM Model

The LSTM model demonstrated notable performance improvements over the baseline logistic regression model, achieving an Average AUC of 0.700 (95% CI: 0.693 - 0.706), compared to the logistic regression’s Average AUC of 0.608 (95% CI: 0.602 - 0.614). Using precision and recall to evaluate the model performance in terms of top 10% high-risk admission patients, LSTM model achieved precision of 0.355 (95% CI: 0.319 – 0.391) and recall of 0.418 (95% CI: 0.389 – 0.448), a significant improvement from the baseline model.

### Features Importance

We utilized permutation importance to further understand the contributions of different features in predicting hospital readmissions within 30 days as shown in Figure 5. Permutation importance measures the decrease in model performance when a single feature’s values are randomly shuffled, effectively breaking the relationship between that feature and the target variable. This method provides an intuitive metric for assessing the impact of each feature on the model’s predictions.

**Figure 5.**
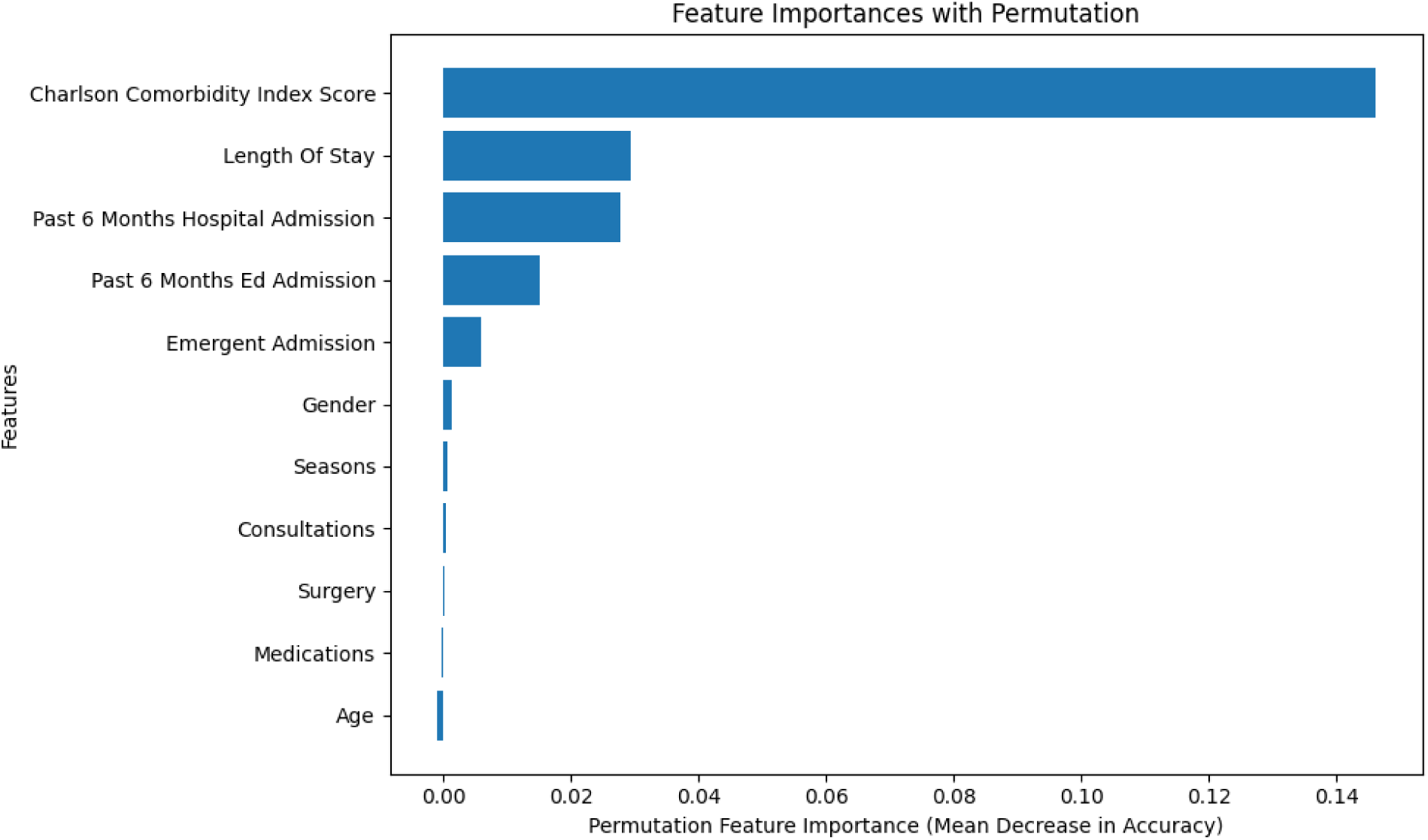
Permutation Analysis

Our analysis reveals that the Charlson Comorbidity Index (CCI) score is the most significant predictor, with an importance value of 0.1424. This finding underscores the substantial influence of chronic conditions on readmission risk. The second most important feature is the length of hospital stay, with an importance value of 0.0313, highlighting the critical role of prolonged hospitalizations in predicting future readmissions.

The number of past six months’ hospital admissions also emerges as a key predictor, with an importance value of 0.0298. This feature indicates that patients with frequent recent hospitalizations are at a higher risk of being readmitted. Additionally, past emergency department admissions contribute significantly, with an importance value of 0.0091.

Interestingly, some features exhibit negative importance values, suggesting they may have a negligible or potentially misleading impact on the model’s predictions. For instance, age has an importance value of -0.0040, and admissions for surgery have an importance value of -0.0007. These negative values indicate that these features, when permuted, might slightly improve the model’s performance, thus questioning their relevance.

Other features, such as season of admission and consultations, show minimal importance values of 0.0010 and 0.0017, respectively, indicating a relatively low impact on the prediction of readmissions. Furthermore, the number of admissions related to medications has an almost negligible importance value of -0.0003.

### Model Adjustment

Recognizing this, we undertook feature selection and engineering to refine our model further. The results are listed in **Table 3**.

**Table 3.**
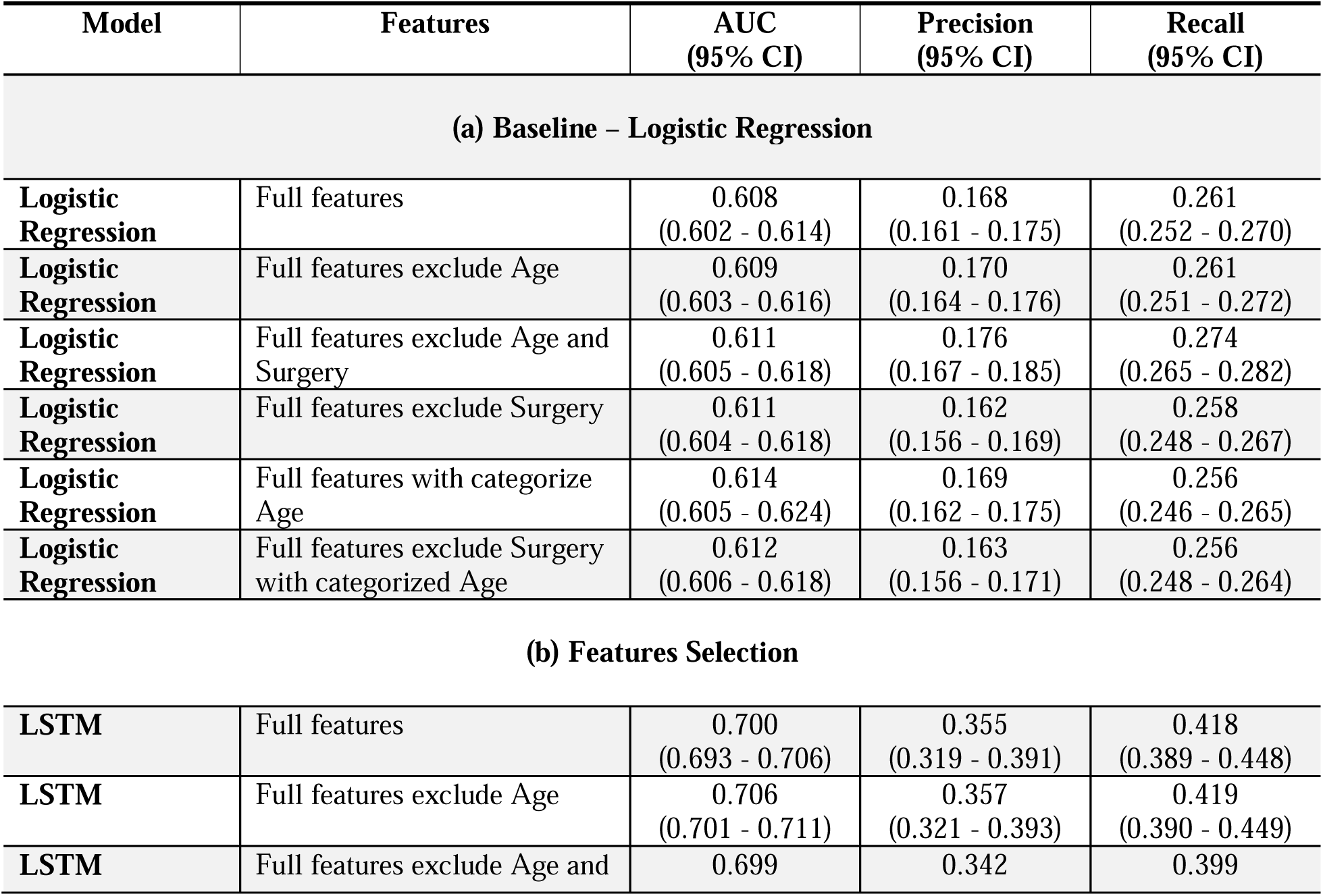
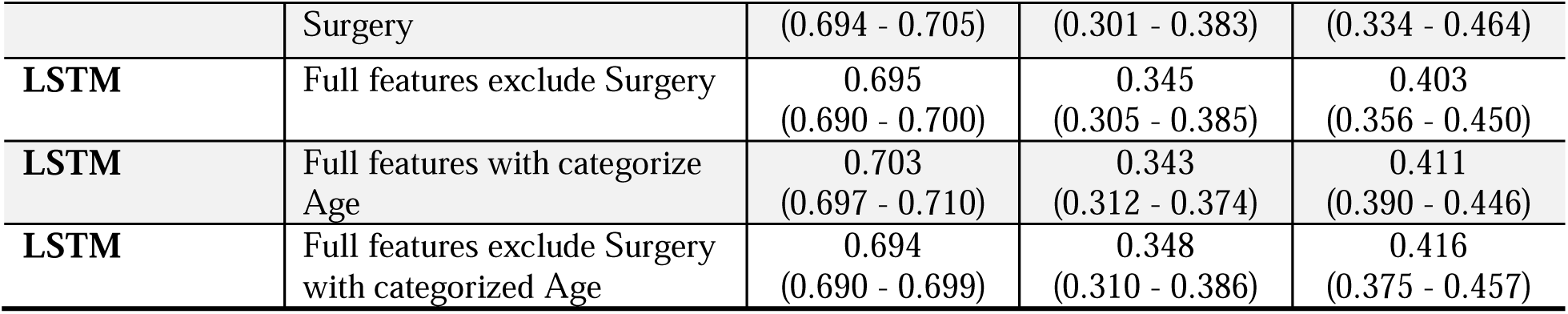
LSTM Model Adjustment.

In our feature selection process, removing the Age feature alone resulted in an Average AUC of 0.706 (95% CI: 0.701 - 0.711), maintaining precision but slightly reducing recall. Removing the Surgery feature led to a decrease in performance with an Average AUC of 0.695 (95% CI: 0.690- 0.700). Combining the removal of both features yielded an Average AUC of 0.699 (95% CI: 0.694 - 0.705), indicating the detrimental impact of excluding these variables together.

Next, we explored feature engineering by categorizing patient ages^44^ into ranges: 65-74, 74-85, and 85+. This categorization improved the model’s Average AUC to 0.703 (95% CI: 0.697 - 0.710), with precision remaining stable and a slight increase in recall. Combining this age categorization with the removal of the Surgery feature resulted in a diminished performance, with an Average AUC of 0.694 (95% CI: 0.690 - 0.699).

Ultimately, our best-performing LSTM model excluded the Age feature but retained the Surgery variable in its original form, achieving an optimal balance between predictive accuracy and recall. These findings highlight the nuanced impacts of feature selection and engineering on model performance, emphasizing the importance of retaining critical variables for robust predictions.

## Discussion

### In-Depth Analysis of Variable Contributions to Readmission Risk: Insights from SHAP

To better understand the impact of each predictor on hospital readmission, we used SHapley Additive exPlanations (SHAP), which is an approach based on cooperative game theory that assigns precise importance values to features in our LSTM model^45^. It is a quantitative measure of the relative influence of each variable on the model’s prediction. The analysis revealed that the Charlson Comorbidity Index (CCI) is the most significant predictor of 30-day hospital readmissions. SHAP values clearly indicate that higher CCI scores are strongly associated with a higher likelihood of readmission. This finding is consistent with the established clinical understanding that patients with multiple comorbidities are at greater risk of adverse outcomes, including readmission. The second-most significant variable after CCI, was hospital length of stay, which could account for the probability of readmissions, likely as a result of the severity of the index admission or complications that delayed recovery.

Contrary to intuition, our analysis also showed an inverse correlation – the more medications and consultations people take in the initial hospital admission, the lower their risk of readmission.

This suggests that better care during the initial hospitalization – including more intensive medical investigations and prophylactic drug treatments – might reduce the risk of patients going home and returning to hospital.

To thoroughly understand the impact of various predictors on hospital readmissions, our analysis employed SHapley Additive exPlanations (SHAP), a robust method rooted in cooperative game theory. SHAP assigns precise importance values to each feature in our LSTM model, allowing us to quantitatively assess the influence of different variables on the model’s predictions. The details of SHAP results are shown in the Figure 6.

**Figure 6.**
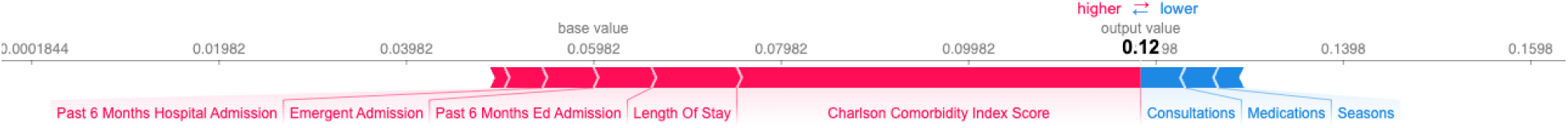
SHAP Force Plot

Another key finding is the gender disparity in readmission rates, with female patients exhibiting a higher propensity for readmission than their male counterparts. This may be due to a variety of factors, including physiological differences, comorbidity profiles, or access to post-discharge care, though further investigation would be required to pinpoint the exact causes.

The model also found that one-time or frequent prior visits to emergency departments over the past six months were moderate predictors of readmission risk, which accords with clinical observations that patients with a history of acute admissions are more likely to experience acute readmission events.

Conversely, other inpatient surgical interventions and seasonal variations demonstrated minimal impact on the readmission prediction, this suggests that while these factors represent only incidental aspects of clinical care.

### Clinical Validation of Model Predictors

To enrich our analysis of the LSTM model’s clinical relevance, we reference pertinent studies that affirm the significance of key model predictors^46^. The Charlson Comorbidity Index (CCI) emerges as a pivotal predictor, corroborated by Dongmei’s research^47^, which delineates a pronounced relationship between high CCI scores and increased short-term readmission risks in heart failure patients, using multivariable logistic regression to pinpoint a risk saturation effect at a CCI score of 2.97 (OR, 2.66; 95% CI, 1.566–4.537). Marco Canepa’s study similarly underscores a direct correlation between elevated CCI scores and heightened readmission in elderly patients^48^.

Jean-Sebastien’s analysis extends the narrative by highlighting the impact of length of stay (LOS) on readmission^49^. Examining 91,723 patients, the study finds that a longer LOS, averaging 6.87 days among readmitted patients versus 5.37 days generally, significantly correlates with increased readmission risk, thus supporting our model’s emphasis on LOS as a substantial predictor.

Prior hospitalizations, especially via emergency departments, and hospitalizations conducted via emergency departments over the past 6 months were independently increased the possibility of unplanned rehospitalization with 30 days of discharge^50^. These factors may account for the total burden of illness, functional status, and social environment^4,51–54^, causing more frequent rehospitalization.

Conversely, the inverse relationship our model identifies between discharge medication counts and readmission likelihood finds empirical backing in Picker’s research, which demonstrates that a higher number of discharge medications is associated with decreased readmission rates, with statistical significance (P < 0.001)^55^.The protective role of consultations is evidenced in a study by David, showing that pharmacist-led medication reconciliations at discharge reduce the 7-day readmission rate significantly from 7.6% to 5.8% in high-risk patients, bolstering our model’s findings^56^.

Gender-specific predictions by our model are supported by research noting physiological differences that influence clinical outcomes^57,58^. Studies highlight that women, with their anatomically smaller body sizes and arteries, are more prone to procedural complications, which is reflected in higher complication rates post-interventions^59^.

The removal of the age variable in our LSTM model adjustment resulted in an improved AUC, precision, and recall, suggesting that age may not be a critical factor in predicting readmission for patients over 65. This observation is supported by research indicating that the impact of age on readmission rates stabilizes beyond this age threshold. Jay’s study reveals that for patients aged over 65, the odds ratio for readmission does not vary significantly with age, highlighting the diminished influence of age on readmission likelihood in this demographic^60^. This finding aligns with the broader clinical understanding that factors other than age might be more predictive of readmission risks in the elderly, guiding more targeted interventions to prevent hospital readmissions.

These studies not only validate the predictive accuracy of our LSTM model but also enrich its clinical interpretability. By anchoring our model’s outputs in robust clinical research, we ensure that the insights it provides are not only scientifically valid but also practically applicable in enhancing patient care and management strategies within healthcare systems. This synthesis of machine learning efficiency and empirical clinical evidence paves the way for targeted interventions that could substantially mitigate readmission rates and improve overall healthcare outcomes.

### Limitations

The limitations of our study are twofold. First, the MIMIC-III database is limited in scope of its single center as it may not represent the wider Medicare population. Although 67.3 million were enrolled in Medicare as of 31 July 2024, we analyzed the data on 21,002 patients. Our particular study might skew the geographical biases and fail to highlight the disparities in hospital readmissions risks depending on geographic variations and racial diversity. In future studies, we plan to overcome these drawbacks by utilizing larger multi-centers^61^ that have a more representative population and add additional demographic factors to our predictive model.

A second limitation was the study’s focus on all-cause hospital readmissions without accounting for specific patients’ chronic disease histories. Although we used the Charlson Comorbidity Index (CCI) to account for comorbidities, this surrogate measure likely fails to adequately capture what we deem to be the ‘chronic’ in older adults with chronic disease. Examples include the multiplicity and complexity involved in the management of a person’s heart failure, diabetes, or some other chronic condition (acknowledging that our heart failure model does better). Future studies could focus on specific chronic diseases to enhance interpretability and inform specific clinical populations, such as the age 65-plus Medicare population. Also, NLP technique can be further utilized to EHR data to provide more valuable features for the prediction^62–64^.

Moreover, this study does not consider external factors which can affect hospital readmission: the external environment or conditions within it such as quality of surrounding areas, distance from the hospital, and patient health behaviors such as smoking or obesity, which can be digitalized from more kinds of data source^65^ . These are vital external factors for seniors and can shed more light on the internal risks of rehospitalization. A possible direction for further research could be to incorporate these external factors and discover their effect on the readmission probabilities while applying the same LSTM deep learning approach to further improve the predictive power and applicability of the model.

## Conclusion

This study underscores the significance of predictive modeling in addressing the challenge of 30- day hospital readmissions among senior Medicare patients. By leveraging a Long Short-Term Memory (LSTM) model, we have demonstrated that incorporating time-series data and focusing on admission-level variables enhances the prediction of readmission risk, surpassing traditional logistic regression models that rely on the LACE index.

The findings highlight that the Charlson Comorbidity Index (CCI) and length of hospital stay are the most influential factors in predicting readmissions. Our LSTM model’s ability to capture temporal dependencies allows it to provide a more nuanced understanding of these predictors compared to baseline models. Moreover, the model’s precision and recall metrics in identifying high-risk patients indicate its potential utility in clinical settings for targeted interventions.

However, the study also reveals limitations related to the data source and scope. The use of a single-center database and the focus on all-cause readmissions may limit the generalizability of the findings. Future research should explore the application of this model across more diverse and larger datasets, as well as its effectiveness in predicting readmissions related to specific chronic conditions.

Overall, this research contributes to the ongoing efforts to reduce hospital readmissions by providing a robust and clinically relevant predictive model. The insights gained from this study can inform healthcare providers and policymakers in developing strategies to improve patient outcomes and reduce healthcare costs. As the healthcare landscape continues to evolve, the integration of advanced machine learning models like the one presented here will be crucial in driving forward data-driven, patient-centered care.

## Permissions

No permission required.

## Data Availability

All data produced in the present work are contained in the manuscript

## Acknowledgments

(See Statements and Declarations page)

## Statements and Declarations

(See Statements and Declarations page)

